# Analysis of CD8 and FOXP3 Expression Ratios in Tumor and Stromal Compartments of Penile Squamous Cell Carcinoma Across Different Subtypes and Grades

**DOI:** 10.1101/2025.01.11.25320379

**Authors:** Sofia Canete-Portillo, Antonio L. Cubilla, George J. Netto, Alcides Chaux

**Affiliations:** The University of Alabama at Birmingham, Birmingham, AL; Instituto de Patologia e Investigación, Asunción, Paraguay; Perelman School of Medicine at the University of Pennsylvania, Philadelphia; Facultad de Medicina, Universidad del Norte, Asunción, Paraguay; Facultad de Ciencias de la Salud, Universidad del Sol, Ciudad del Este, Paraguay

**Keywords:** Penile Squamous Cell Carcinoma, CD8+ T Cells, FOXP3+ Regulatory T Cells, Immunohistochemistry, Tumor Microenvironment

## Abstract

**Background:** Penile squamous cell carcinoma (PSCC) remains a relatively rare but formidable malignancy, especially in regions with limited access to preventive measures. The tumor microenvironment (TME) –specifically, the balance between CD8+ cytotoxic T cells (CTLs) and FOXP3+ regulatory T cells (Tregs)– has emerged as a pivotal determinant of tumor progression and immune evasion. This study aimed to evaluate CD8+/FOXP3+ ratios in both tumor and stromal compartments across different PSCC subtypes and grades.

**Methods:** This retrospective study analyzed tissue microarray (TMA) samples from 108 patients with invasive PSCC. Immunohistochemical staining for CD8+ and FOXP3+ was performed. Tumor and stromal compartments were assessed separately. Ratios of CD8+/FOXP3+ were categorized as CD8 > FOXP3 or CD8 ≤ FOXP3. Associations with histologic subtype and grade were examined using Chi-Square or Fisher’s Exact tests, with Cramér’s V indicating effect size.

**Results:** Eighty TMA spots (15.2% of the total) had quantifiable data for both markers. We observed a significant association between CD8+/FOXP3+ ratio and histologic grade in both tumor (P=0.03) and stromal compartments (P=0.02), with moderate effect sizes (Cramér’s V ~ 0.3). Although no statistically significant associations emerged for histologic subtype, effect size measures suggested potential immune-infiltration differences across subtypes. Descriptive analyses indicated that tumor compartments often contained fewer T cells overall, while stromal areas demonstrated robust infiltration patterns.

**Conclusions:** Tumor grade appears to influence the relative infiltration of cytotoxic and regulatory T cells in PSCC, underscoring the need for compartment-specific immune profiling. These observations highlight the potential utility of CD8+/FOXP3+ ratios as prognostic markers and in guiding future immunotherapeutic strategies. Prospective studies incorporating larger cohorts and HPV stratification could further clarify the immunobiology of PSCC and inform personalized treatment approaches.

## INTRODUCTION

Penile squamous cell carcinoma (PSCC) is a relatively uncommon malignancy, yet it presents a significant challenge in oncology due to its aggressive nature and the substantial morbidity and mortality it inflicts, particularly in regions with limited access to healthcare and preventive strategies (Chaux et al., 2013; Júnior et al., 2022). Despite its rarity, PSCC’s impact on patients’ quality of life, combined with limited therapeutic options, makes it a pressing area of research. The tumor microenvironment (TME), a complex ecosystem encompassing immune cells, stromal cells, and extracellular matrix components, plays a key role in cancer progression and response to therapy, thus making it crucial to explore for PSCC (Vries et al., 2019).

Within the complex TME, the interplay between CD8+ cytotoxic T cells (CTLs) and FOXP3+ regulatory T cells (Tregs) is pivotal in determining the outcome of the immune response. CD8+ CTLs are essential effector cells in adaptive immunity, capable of recognizing and eliminating tumor cells (Ottenhof et al., 2018). Conversely, FOXP3+ Tregs are a subset of immunosuppressive T cells that can inhibit the activation and function of CTLs, thereby promoting immune evasion and tumor progression (Mumba, 2024). The balance between these opposing forces is key, and a shift towards an immunosuppressive environment can facilitate tumor growth and metastasis.

CTLs are the primary effectors of cell-mediated immunity, tasked with directly recognizing and eliminating tumor cells expressing specific antigens via the MHC-I pathway (Ottenhof et al., 2018). Their ability to infiltrate tumor nests and exert cytotoxic functions are critical factors in controlling tumor growth. Conversely, Tregs play a role in maintaining immune homeostasis, preventing excessive immune responses, and maintaining self-tolerance. However, within the TME, Tregs can suppress the activity of CTLs, facilitating immune evasion and tumor progression, therefore shifting the equilibrium towards tumor progression (Hladek et al., 2022; Mumba, 2024).

Previous studies in PSCC have explored the role of other immune markers, such as programmed death-ligand 1 (PD-L1), and its correlations with CD8+ and FOXP3+ cells (Cocks et al., 2017; Montella et al., 2022; Udager et al., 2016). These studies have provided valuable insights into the immune microenvironment, identifying potential pathways of immune resistance. However, understanding the balance between cytotoxic and regulatory T cell populations, expressed through the CD8/FOXP3 ratio, is also crucial for a comprehensive characterization of the TME.

Therefore, this study aims to characterize the ratio of CD8+ cytotoxic T cells to FOXP3+ regulatory T cells within both the tumor and stromal compartments of PSCC, considering different histological subtypes and grades. To achieve these aims, we performed an immunohistochemical analysis to quantify CD8+ and FOXP3+ cells in a tissue microarray of PSCC samples, classifying them according to the tumor’s histological subtype and grade, and evaluating the distribution of the CD8/FOXP3 ratio within tumor and stromal compartments. We then used statistical analyses to correlate the distribution of the CD8/FOXP3 ratio with the histological features.

## MATERIALS AND METHODS

### Case Selection and Tissue Microarray Construction

The study analyzed tissue samples from 108 patients with invasive squamous cell carcinoma of the penis from the consultation files of one of the authors (ALC). Cases were selected based on availability of formalin-fixed, paraffin-embedded tissue blocks, with 1–4 blocks selected from each case to ensure adequate representation of tumor heterogeneity. Four tissue microarrays (TMA) were constructed at the Johns Hopkins TMA Lab Core (Baltimore, MD) using established procedures (Fedor & De Marzo, 2005). Three tissue cores of 1 mm each were obtained per block, providing 3–12 spots per case. Normal tissue from various anatomical sites were included as control tissue. A total of 528 TMA spots were evaluated from the 108 cases.

### Morphological Evaluation

Pathologic features were evaluated using H&E-stained tissue sections by three expert genitourinary pathologists (ALC, GN, AC). Histologic subtyping was performed on whole tissue sections using the 5th edition (2022) WHO criteria for classification of tumors of the urinary system and male genital organs (Moch et al., 2022). Histologic grading was conducted spot by spot using previously published and validated criteria (Chaux, 2015). Grade 1 tumors showed well-differentiated cells with minimal basal/parabasal cell atypia. Grade 3 tumors demonstrated anaplastic cells with nuclear pleomorphism, coarse chromatin, prominent nucleoli, irregular and thickened nuclear membranes, and abundant atypical mitoses. Grade 2 tumors represented an intermediate category, not fitting criteria for grades 1 or 3.

### Immunohistochemical Analysis

Immunohistochemical (IHC) staining was performed on the TMA sections using a Ventana Benchmark XT automated immunostainer (Ventana Medical Systems, Tucson, AZ, USA). The sections were deparaffinized, rehydrated, and then subjected to heat-induced epitope retrieval using CC1 buffer (Ventana Medical Systems). For the detection of CD8+ cytotoxic T lymphocytes, a mouse monoclonal antibody against CD8 (clone C8/144B, Dako, Carpinteria, CA, USA) was used at a dilution of 1:100. For the detection of FOXP3+ regulatory T cells, a mouse monoclonal antibody against FOXP3 (clone 236A/E7, Abcam, Cambridge, UK) was used at a dilution of 1:200. The sections were incubated with the primary antibodies for 30 minutes at room temperature. Detection was performed using the Ventana OptiView DAB IHC Detection Kit, according to the manufacturer’s protocol, followed by counterstaining with hematoxylin. Sections were then dehydrated, cleared, and mounted with permanent mounting media.

### Quantification of Immunostaining

The quantification of CD8+ and FOXP3+ immunostaining was performed manually by two independent, experienced pathologists (SCP, GJN), who were blinded to each other’s results, and to the clinicopathological data. The quantification was performed separately for the tumor and stromal compartments within each tissue core. In each compartment, the number of CD8+ and FOXP3+ positive lymphocytes were counted in at least 10 high-power fields (400x magnification), with a consensus agreement if disagreements occurred. The CD8+/FOXP3+ ratio was then classified into two categories for each sample: CD8 ≤ FOXP3 and CD8 > FOXP3, based on the number of positive cells in each high-power field. The intensity of the immunostaining in both cellular compartments, was categorized as absent (no staining), weak, moderate, or strong.

### Statistical Analysis

Statistical analyses were conducted using Python version 3.12.7, within the Visual Studio Code environment, leveraging the pandas library for data manipulation and the scipy.stats module for statistical calculations. Specifically, the association between the categorized CD8+/FOXP3+ ratio (CD8 ≤ FOXP3 and CD8 > FOXP3) and the histological subtype and grade within both the tumor and stromal compartments was assessed using the Chi-Square test of independence. In cases where the expected cell count in any cell was under 5, a Fisher’s Exact test was used. The Chi-square test, and Fisher’s Exact test when applicable, were used to evaluate the statistical independence of the T-cell ratios concerning the categorical variables, where a P-value of less than 0.05 was considered statistically significant.

Additionally, to quantify the strength of any statistically significant association, we calculated Cramér’s V, an effect size measure appropriate for categorical data. Cramér’s V values were interpreted as follows: around 0.1 indicating a small association, around 0.3 indicating a moderate association, and around 0.5 indicating a strong association. No corrections for multiple comparisons were made, as our intention was to identify potential differences, which could be further evaluated in future studies.

## RESULTS

Eighty samples (15.2%) showed quantifiable data for both CD8 and FOXP3 expression. Of these, 32 (40.0%) were classified as the usual subtype, 18 (22.5%) as the warty subtype, 15 (18.8%) as the basaloid subtype, and 15 (18.8%) as the warty-basaloid subtype. Regarding the histological grade, 16 (20.0%) tumors were classified as grade 1 (well-differentiated), 40 (50.0%) as grade 2 (moderately differentiated), and 24 (30.0%) as grade 3 (poorly differentiated).

### CD8+/FOXP3+ Ratios in Tumor and Stromal Compartments

The distribution of CD8+/FOXP3+ ratios within the tumor and stromal compartments of PSCC samples, considering different histological subtypes and grades, is shown in Table 1.

A Chi-Square test of independence was used to analyze the association between the CD8+/FOXP3+ ratio and the histological subtype in the tumor compartment. The test revealed no statistically significant association (χ^2^(3) = 7.30, p = 0.06) and a Cramér’s V of 0.30, suggesting a moderate association. The association between the CD8+/FOXP3+ ratio and the tumor grade in the tumor compartment was also analyzed using a Chi-Square test, where a significant association was found (χ^2^(2) = 6.92, p = 0.03) and a Cramér’s V of 0.29, which suggests a moderate association as well.

In the stromal compartment, the Chi-Square test for the association between the CD8+/FOXP3+ ratio and the histological subtype also revealed no statistically significant association (χ^2^(3) = 6.42, p = 0.09) with a Cramér’s V of 0.28, indicating a small association. The association between the CD8+/FOXP3+ ratio and the tumor grade in the stromal compartment showed a statistically significant association (χ^2^(2) = 7.19, p = 0.02) and a Cramér’s V of 0.30, indicating a moderate association.

In addition to classifying each TMA spot categorically as “CD8 > FOXP3” or “CD8 ≤ FOXP3,” we calculated basic descriptive statistics for the raw CD8+ and FOXP3+ cell counts in both the tumor and stromal compartments. Overall, the median (IQR) number of intratumoral CD8+ cells was 6 (0–28) per high-power field (HPF), while intratumoral FOXP3+ cells showed a median (IQR) of 2 (0–8) per HPF. In the stromal compartment, the median (IQR) CD8+ cell count was 30 (10–50) per HPF and the median (IQR) FOXP3+ cell count was 20 (5–40) per HPF. Although our principal focus was on the CD8+/FOXP3+ ratio, these descriptive statistics provide additional context regarding the overall density of T-cell infiltration.

## DISCUSSION

One of the primary aims of this study was to assess the spatial distribution and balance between CD8+ cytotoxic T cells and FOXP3+ regulatory T cells across different histological subtypes and grades of PSCC. This focus on the CD8+/FOXP3+ ratio stems from the hypothesis that a higher prevalence of cytotoxic T cells relative to regulatory T cells may correlate with more robust antitumor responses (Chakraborty et al., 2018; Kim et al., 2019). In line with prior investigations of immune cell infiltration in other squamous cell carcinomas, our analysis revealed moderate associations between the CD8+/FOXP3+ ratio and histologic grade in both tumor and stromal compartments, implying that tumor differentiation level exerts a measurable influence on local immune dynamics (Lohneis et al., 2014; Wansom et al., 2011).

Notably, although the correlations with subtype did not reach conventional statistical significance, the effect size measures suggest potentially meaningful distinctions in immune cell infiltration across different histologies—an observation that could become more pronounced with larger sample sizes or more granular subtyping (Hladek et al., 2022). These findings underscore the complexity and heterogeneity of immune responses in PSCC. They also highlight the importance of evaluating both tumor and stromal compartments separately, given that immune infiltration patterns and their prognostic significance can differ substantially depending on the microanatomical context (Mezheyeuski et al., 2018).

Our findings on the moderate association between the CD8+/FOXP3+ ratio and histologic grade align with observations in other squamous cell carcinomas. For instance, Kim et al. (2019) reported that a lower proportion of regulatory T cells relative to cytotoxic T cells was associated with improved therapeutic responses in non-small cell lung cancer. Similarly, in head and neck squamous cell carcinoma, higher densities of tumor-infiltrating CD8+ T cells have been linked to enhanced survival (Wansom et al., 2011). These studies suggest that the predominance of cytotoxic T cells may tip the immunological balance toward a more effective antitumor response—an idea mirrored in our PSCC population insofar as higher-grade tumors exhibited significantly different CD8+/FOXP3+ ratios.

It is worth noting that, although the association of CD8+/FOXP3+ ratios with histologic subtype did not achieve strict statistical significance in our cohort, similar trends have been observed in other tumor types concerning variations in immune infiltration by subtype (Lukešová et al., 2014). This may indicate that, in PSCC, other microenvironmental factors—such as HPV status or local cytokine profiles—could be modulating immune cell migration and function, thereby overshadowing subtype-specific differences (Mumba, 2024). Moreover, the consistent finding of moderate effect sizes (Cramér’s V ≈ 0.3) in both tumor and stromal compartments corroborates previous literature emphasizing that immune responses can differ between these two microanatomical areas (Mezheyeuski et al., 2018). This nuance underscores the importance of compartmental analyses when investigating the immune microenvironment, since T cells within the tumor nests themselves may exert direct cytotoxic effects, while stromal T cells might be subject to different immunomodulatory signals (Chakraborty et al., 2018).

The interplay between CD8+ CTLs and FOXP3+ regulatory Tregs in PSCC likely operates through multiple immunological pathways. On one hand, CD8+ T cells are central immune effectors that recognize tumor-associated antigens via the major histocompatibility complex (MHC) class I pathway, leading to the targeted elimination of neoplastic cells (Chakraborty et al., 2018). Their functionality, however, can be curtailed by an abundance of Tregs, which are known to use diverse mechanisms to maintain immune homeostasis—ranging from the secretion of suppressive cytokines (e.g., IL-10, TGF-β) to direct cell-contact inhibition mediated via checkpoint molecules (Sharma et al., 2019). Within the context of PSCC, heightened Treg infiltration may support an immunosuppressive milieu that hampers effective cytotoxic responses (Wang et al., 2020).

Tumor grade itself could further modulate this interplay. Poorly differentiated tumors often display heightened genomic instability and increased neoantigen load, features that can theoretically elicit greater T cell infiltration (Kim et al., 2019). Concomitantly, however, such tumors may foster the recruitment or expansion of immunosuppressive T cell populations, including Tregs, through the release of specific chemokines or induction of tolerogenic dendritic cells (Chahoud et al., 2020). This dynamic may explain why we observed moderate associations—rather than uniform patterns—between the CD8+/FOXP3+ ratio and tumor grade in both the tumor and stromal compartments.

Additionally, emerging evidence suggests that the human papillomavirus (HPV) status of PSCC may affect T cell recruitment and phenotype, possibly by providing viral antigens that stimulate a stronger Th1-type immune response (Lohneis et al., 2014; Mumba, 2024). Our study did not specifically address HPV status, but prior work implies that HPV-positive PSCCs could harbor more robust CD8+ T cell infiltrates relative to HPV-negative tumors, potentially influencing the net balance of cytotoxic versus regulatory T cells (Cipollini et al., 2021). Future efforts incorporating HPV stratification may reveal more nuanced mechanisms by which viral oncogenes shape the immune landscape of PSCC.

An enhanced understanding of the balance between cytotoxic (CD8+) and regulatory (FOXP3+) T cells within PSCC has several potential applications in clinical practice. First, quantifying the CD8+/FOXP3+ ratio could help refine prognostic models, given the emerging evidence that immune cell infiltration patterns can correlate with outcomes in various squamous cell carcinomas (Kim et al., 2019; Montella et al., 2022). Specifically, a higher density of cytotoxic T cells relative to Tregs may reflect a more active antitumor response, possibly signaling improved survival or reduced metastatic propensity in PSCC.

Second, immune-based therapeutic strategies could benefit from these findings. Investigational approaches targeting immune checkpoints—such as PD-1/PD-L1 inhibitors— have begun to show promise in advanced or refractory scenarios of genitourinary cancers (Chahoud et al., 2020; Udager et al., 2016). While these agents primarily aim to reinvigorate T cell function, they may be complemented by strategies to mitigate excessive Treg-related immunosuppression, such as selective Treg depletion or blockade of Treg-supporting pathways (Miller et al., 2015). Integrating analysis of the CD8+/FOXP3+ ratio into clinical trials could help identify patients most likely to respond to immunotherapies, thereby optimizing patient selection and potentially improving outcomes.

Third, the tumor-versus-stroma compartment analysis underscores the need for careful microanatomical evaluation when interpreting immunohistochemical or molecular data (Mezheyeuski et al., 2018). In practice, this might entail using multiplex immunofluorescence or other advanced imaging modalities in the diagnostic setting to delineate the precise location of immune cells. Such detailed data may yield insights into the functional interplay between T cells and tumor cells, informing both prognostic assessments and therapeutic approaches.

Despite our findings, several limitations should be acknowledged. First, the retrospective nature of the study introduces potential biases in case selection and data availability (Vries et al., 2019). Although we endeavored to maintain consistency across patient materials and immunohistochemical protocols, unrecognized differences in tissue quality or fixation methods may influence results. Second, the relatively modest sample size—despite including up to 108 PSCC tissues—reduces the power to detect more subtle associations, particularly regarding distinct histological subtypes. This limitation is reflected in the lack of statistically significant findings for certain subgroup comparisons, suggesting that a larger cohort may be necessary to clarify subtype-specific immune patterns (Mumba, 2024). Third, although the compartmental analysis of tumor versus stroma provides a more nuanced view of the immune landscape, it remains a snapshot in time. We did not evaluate longitudinal changes in immune cell populations, nor did we incorporate functional assays (e.g., cytokine profiling, T cell receptor repertoire analyses) that could elucidate the biological underpinnings of these immune interactions (Mezheyeuski et al., 2018).

Lastly, we did not stratify cases by human papillomavirus (HPV) status, despite accumulating evidence that the presence of oncogenic HPV can influence immune responses in PSCC (Chipollini et al., 2021). Future investigations that integrate HPV typing and additional immunological markers could yield deeper insights, particularly into how viral oncogenes shape the CD8+/FOXP3+ balance (Lohneis et al., 2014).

To build on these findings, prospective studies incorporating larger cohorts are essential to refine and validate the prognostic value of CD8+/FOXP3+ ratios in PSCC. Stratifying patients by HPV status and employing advanced techniques—such as multiplex immunofluorescence, spatial transcriptomics, or single-cell RNA sequencing—could yield a more comprehensive understanding of how these immune cell populations interact and evolve over time (Mei et al., 2022). Functional assays examining cytokine production or T cell receptor clonality may also shed light on the biological pathways underpinning the observed immunologic patterns (Chakraborty et al., 2018). Moreover, clinical trials that integrate CD8+/FOXP3+ ratio analyses into patient selection criteria for immunotherapeutics might further delineate subsets likely to respond to checkpoint inhibitors or novel Treg-modulating agents (Chahoud et al., 2020; Miller et al., 2015).

In conclusion, our study highlights the importance of evaluating both tumor and stromal compartments when assessing the CD8+/FOXP3+ ratio in PSCC. The moderate associations we observed between this ratio and histologic grade underscore the complexity of the tumor immune microenvironment, suggesting that divergent differentiation states may differentially recruit or sustain cytotoxic versus regulatory T cells. While these observations warrant further exploration in larger, prospectively collected cohorts, they lay the groundwork for integrating immune profiling into prognostic models and guiding personalized immunotherapeutic strategies. By combining robust immunophenotypic assessments with advanced molecular analyses, future research has the potential to refine our understanding of PSCC pathobiology and improve outcomes for affected patients.

## Data Availability

We have made the entire dataset used openly accessible to the scientific community. Interested researchers and collaborators can access this resource through the following link: https://doi.org/10.6084/m9.figshare.28110671.v1.

