## Supplementary material for "Analysis of CD8 and FOXP3 Expression Ratios in Tumor and Stromal Compartments of Penile Squamous Cell Carcinoma Across Different Subtypes and Grades": Table 1

**Table 1:** CD8+/FOXP3+ Ratios in Tumor and Stromal Compartments

| Feature | N (%) | Tumor compartment (%) | | Stromal compartment (%) | |
|---|---|---|---|---|---|
| | | CD8 > FOXP3 | CD8 ≤ FOXP3 | CD8 > FOXP3 | CD8 ≤ FOXP3 |
| Histological Subtype | | | | | |
| Usual | 32 (40.0) | 21 (65.6%) | 11 (34.4%) | 17 (53.1%) | 15 (46.9%) |
| Warty | 18 (22.5) | 9 (50.0%) | 9 (50.0%) | 12 (66.7%) | 6 (33.3%) |
| Basaloid | 15 (18.8) | 5 (33.3%) | 10 (66.7%) | 4 (26.7%) | 11 (73.3%) |
| Warty-Basaloid | 15 (18.8) | 6 (40.0%) | 9 (60.0%) | 5 (33.3%) | 10 (66.7%) |
| Histological Grade | | | | | |
| Grade 1 | 16 (20.0) | 12 (75.0%) | 4 (25.0%) | 13 (81.3%) | 3 (18.8%) |
| Grade 2 | 40 (50) | 20 (50.0%) | 20 (50.0%) | 24 (60.0%) | 16 (40.0%) |
| Grade 33 | 24 (30.0) | 9 (37.5%) | 15 (62.5%) | 11 (45.8%) | 13 (54.2%) |
